# Optic Disc and Vessel Segmentation from Fundus Image using Miniunet Architecture

**DOI:** 10.1101/2024.12.28.24319728

**Authors:** Subrata Jana, Paramita Sarkar

## Abstract

The segmentation of the retinal blood vessels significantly impacts the early diagnosis of conditions that affect the eyes, such as diabetes and glaucoma. This work uses fundus pictures to segment retinal blood vessels using mini-Unet algorithms. Many thousands of gloss training samples are required for effective deep network training, which is highly permissible. Mini-Unet starts with a spatial concentration element that multiplies the concentration map by the input feature for adaptive element enhancement and deduces the concentration map in addition to the spatial measurement. Mini-unit architecture creates a structure of a constricting path that enables a particular position. We design three types of primal action, such as Unet_Gatting, AttnGating Block, and convolution, for image segmentation. The mini_unet model is a modified version of the unet model. In Figure 1.1, mention where to modify my techniques. The success of the planned network structure was verified by two segmentation responsibilities: retina vessel segmentation and lung segmentation. Our technique used segmentation of existing DRIVE and STARE datasets. In fundus images using deep learning-based convolutional neural networks. In our technique, accuracy is .95337.

## 1. Introduction

Nowadays, 450 million people worldwide are diabetics, according to WHO statistics. The most clinically vital peril factors for succession to vision loss consist of the duration of diabetes, hyperglycemia, and hypertension. Controls of serum glucose and blood pressure are successful in avoiding vision loss due to diabetic retinopathy. Glaucoma [63] and retinopathy can have serious effects on diabetics. Arteriolosclerosis, diabetes, and retinopathy are common retinal diseases as well as systematic diseases. Retinal disease occurs when blood pressure is high. Eye vessel structures are tortuous and have many small branches. Pathological changes cannot afford perfect information but also help to classify the disease’s brutality and involuntarily analyze the disease.

The retinal test has been widely used worldwide and is non-invasive and reasonably priced to administer. Giving doctors accurate interpretations of medical images is the main objective of the Computer-Aided Diagnosis (CAD) method, which aims to improve care for many patients. Deep learning networks generally received that unbeaten training of deep networks is mandatory for thousands of interpretable training samples. One main complexity in the eye vessel segmentation mission is that those eye vessels have no significance dissimilar in manifestation from the background, especially for the micro eye vessels in the noisy image.

Image segmentation [32] means partitioning individual objects and classifying pixel problems with semantic segmentation [39,50]. Semantic segmentation makes labeling pixel-level groupings of item categories for every pixel in a picture possible. Classifying single-label entire images is an extremely challenging task. By recognizing and elucidating each object of concern in the image, segmentation significantly broadens the options for semantic segmentation.

A deep learning network is used to classify the task and where the particular network label is to a specified input image. Convolution function, various optimization algorithms, activation function, and dropout were among the structural aspects this network employed [40] as a regularizer. Numerous training data sets and many network parameters were required for this network. Instead of marking the image level, each pixel must be labeled in biomedical image segmentation [54].

In deep learning networks, the first fully convolutional neural network (FCN) [15, 18] for image segmentation. U-Net [16] is the extended architecture that requires a large number of training data to accomplish good segmentation results then. A constructing path and an expanding path are combined in this network. While building, many common maps with condensed dimensions are removed from the input data. The stage up-convolution uses the expanding path to create a segmentation map.

**FIGURE 1.1.**
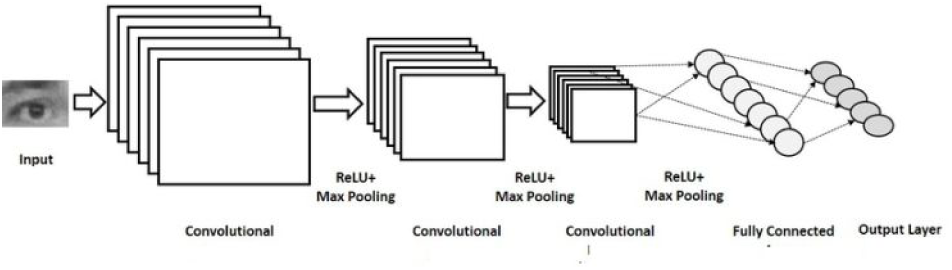
Architecture of neural networks with convolutions.

The Convolutional Neural Network (CNN) consists of three primary layers: convolution, pooling, and drop-off layers, which are the first three layers. i) The first convolution layer uses extracted features with weight input data using the kernel. ii) Most layer types, including convolution, recurrent, and dense linked layers, use the second layer, also referred to as the Dropout layer. The dropout layer implemented the visible input layer and all hidden layers. Iii) The pooling layer comes in at number three.

The pooling layer minimizes the height and width information by maintaining a constant number of channels in the input matrix. One step to lessen the complexity is the computation. CNN’s architecture: MobileNet [34, 48], DenseNet [38], ReluNet [27], and AlexNet [8]. Google LeNet [12], VggNet [14].Low-level characteristics like edge, slope, shape, particular item, and color are eliminated from input images via convolutional neural networks. Following the convolution layer in the Unet model comes a new layer called the pooling layer. It is always about 2x2 pixels with a two-pixel step. This means that the pooling layer always reduces the size of each feature map by a factor of two.

Segmenting retinal vessels and optic discs on the DRIVE and STARE datasets is the primary objective of this work. The input layer of the first, which features a three-level Mini-Unit, contains sixteen filters. Since the anticipated speed increase is not seen, we want to improve its performance by including common deep-learning blocks in the architecture. Five-layer architecture is the fundamental U-Net structure; we suggested a four-layer design in the mini_Unet Model. We created an experiment to compress the fundamental Unet structure to test the theory. The convolutional layer— pooling and dropout—is decreased by the number of levels, convolutional levels per layer, and filters. After execution, lower superpixels to find vessels and delete any unneeded regions. This technique has better results than existing techniques and accurate optic disc location and vessel Segmentation.

The term “conventional segmentation” refers to dividing a picture into several segments, which could be collections of pixels or visual objects.

Image histograms, thresholds, edge detection, and other clustering methods all make use of segmentation algorithms. Although this kind of segmentation is quick and simple, its accuracy is not very high. Nowadays, more researchers use deep learning architectures in computer vision, such as Generative Adversarial Networks (GANs) [10], encoder-decoder [21], AlexNet [8], VGGNet [14], GoogLeNet [12], ISqEUnet [55], OCT [92], Recurrent Neural Networks (RNN) [2], Long-Short-Term Memory (LSTM) [4], and Conventional Neural Networks (CNN) [5, 33].

This paper is divided into six parts. The methods for extracting blood vessels that are currently in use are described in Section 2. The architecture, algorithm, and suggested method are explained in Sections 3, 4, and 5. The experimental results are described in Section 6. The conclusion is explained in Section 7.

## 2. PURPOSE OF THE REVIEW

We aim to identify glaucoma by applying feature extraction and classification methods, which are fundamental research problems in this area. A comparison list of previous studies in this field, complete with comparative analysis, has been supplied in the given table. The following lists various diagrammatic representation techniques.

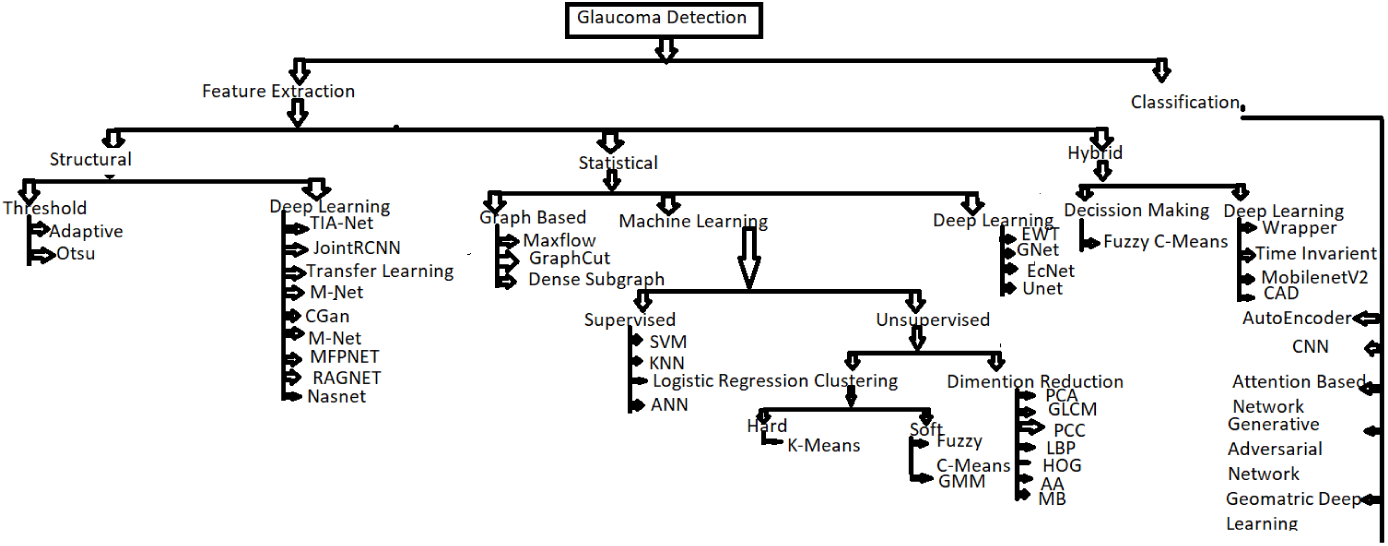

### 2.1. Feature Extraction

To convert raw data into a format that is easier to handle and conducive to analysis, the feature extraction procedure is essential. It entails extracting a collection of representative characteristics from the input data, which form the basis of further analytical procedures. Three steps make up the feature extraction process: three types of features: structural, statistical, and hybrid. These categories fit into the overall pipeline for glaucoma identification, where hybrid features incorporate both structural and statistical features, statistical features give information acquired from images, and structural features record the physical aspects of the optic disc and vascular segmentation. To improve the model’s accuracy and dependability, the extracted informative characteristics are then used as inputs for training.

#### 2.1.1. Structural Feature

There are two kinds of structural features. One is based on thresholds, and the other is deep learning. For doctors to assess retinal structures related to glaucoma, structural imaging measures that concentrate on the physical properties of the optic nerve head are crucial. The accuracy of deep learning algorithms can be greatly increased by incorporating clinical information. These metrics provide objective information that can be used to monitor and assess glaucoma detection. Image segmentation was used by the threshold-based approach, which produces a binary image with a white pixel indicating an optic disc. Clinicians frequently employ the CDR, a crucial structural measurement that can be extracted from fundus images, because it is relevant and vital in the assessment of glaucoma. A. Isaac, M. Parthasarathi, et al. (2015)[19] suggested an adaptive threshold framework for OC and OD segmentation using geometrical features. The program harvests and monitors blood vessels in the Disc Region, determining the initial ending location of various arteries and connecting them to delineate the contours of the OC. It uses employee diagnostic procedures similar to those employed by ophthalmologists. To categorize the fundus image, they also calculated the vertical CDR, which is the distance between the upper and lower locations of the OC and OD.

In deep networks, it is employed. It has routes for both decoding and expanding. By using the Squeeze Expanded module frequently, the contracting path eliminates higher-level features. To create a superposition of global and local in sequence, ISqEUNet [55] combines the contextual in sequence from the up-sampling path with the local in sequence from the down-sampling path. It works well for segmenting irises. This network improves segmentation accuracy while lowering the number of trainable parameters.

#### 2.1.2. Deep Learning

Deep learning models are used to detect minor changes that are suggestive of glaucoma and to identify complex disease features and progression criteria. We investigate the relationship between structural function and the evolution of glaucoma and forecast functional impairment based on structural degradation of the eye. several methods, including M-Net[65], cGAN[44], Tia-Net[64], RagNet[81], and NasNet[66].

RNN [2, 24, 25] is the only neural network type having internal memory, making it a strong and resilient network that fits into the most promising algorithms now in use. A recurrent neural network (RNN) is one type of neural network that can be used to simulate sequences. Its feed-forward network behavior is similar to that of the human brain.

Recurrent neural networks are extended by long short-term memory (LSTM) networks [4, 26,51]. RNNs can store inputs for extended periods because of LSTMs. This makes it possible for LSTMs to store information in a memory, such as computer memory. Based on time series data, LSTM networks are ideal for classification, processing, and forecasting because there may be lags of unknown duration between important events.

Among other deep learning methods, it uses convolutional neural networks [5,26]. For the model to provide output that was extracted from the original data sets, machine learning— which can be either supervised or unsupervised [17,19]—aims to automatically identify and learn patterns in incoming data. For issues like image-to-image translation, GANs offer a route to advanced domain-specific data augmentation and generative solutions.

The encoder-decoder model is one way to use recurrent neural networks for sequence-to-sequence forecasting [21, 41]. It was first created for machine translation problems, but it has shown success in the related sequence-to-sequence prediction tasks. Among other things, chatbots, text summarization, and question answering use these modes.

While deep learning models can be constructed using quantity-labeled training data from scrapes on new applications or databases, there is sometimes not enough labeled data available to train a model from a scrape. One can use relocation knowledge to solve this problem. To categorize photos for tasks like object detection, texture classification, and face recognition, for example, a model trained on ImageNet [16] was employed [27,35,42,49]. 6].ImageNet provides a large number of databases that can be used for object localization tasks. The pre-trained model should be able to confine the semantics in a sequence of the image requisite to the segment.

Deep learning based on segmentation-based techniques is good at getting accurate CDR measurements. However, issues like overlapping regions, low contrast between regions, homogeneity of the OD shape, insufficient labels, and mistakes in the intermediate step still cause problems.

The semi-supervised learning model used in Zhao, Chen, Liu, et al. (2020) [69]’s proposed method for direct CDR estimation eliminates the need for a segmentation step in between. This method is structured in two cascaded ways: first, it represents the unsupervised features of fundus images using a convolutional neural network (CNN); second, it predicts the CDR value with an accuracy of 90.50 percent using a random forest regressor.

One architecture that is frequently used in the field of computer vision deep learning is CNN. K. Fukushima developed the first CNN model, a self-organizing neural network model for a pattern recognition process independent of positional changes in the publication Neocognitron [1]. Figure 1.1 shows the CNN architecture, which was developed and presented by A. Waibel et al. [3] and Y. Lecun et al. [5].

Raja, Hassan, and Akra (2021) [67] presented a framework and methodology that includes a hybrid convolutional network that extracts the ganglion cell complex region and retinal nerve fiber layer, enabling quantitative screening of glaucomatous subjects to extract thickness-related information. To determine the degree of glaucoma, they employed a support vector machine, which produced an accuracy of 91.17 for AIFO data sets.

A deep learning model framework utilizing the NasNet architecture, initially trained on ImageNet, was proposed by Lee, Kim, Park, et al. (2020) [57]. The tailback feature on the SD-OCT images was extracted using all of the NasNet layers that had been frozen. The RNFL thickness map, RNFL deviation map, GCIPL thickness map, and GCIPL deviation map characteristics are extracted by the NasNet architecture. This model has a sensitivity of 94.7% and an accuracy of 99%. Deep learning has a high sensitivity and specificity for detecting structural alterations associated with glaucoma.

An end-to-end region-based convolutional neural network joint optic disc and cup segmentation was proposed by Jiang, Duan, et al. (2020) [58]. The JointRCNN model suggested a Disk Proposal Network (DPN) and a Cup Proposal Network (CPN) to generate bounding box optic discs, which are first chosen and then proceed to be forward propagated as the foundation for optic cup detection. The JointRCNN model outperforms state-of-the-art techniques for glaucoma detection and optic disc and cup segmentation tasks. This technique improves the accuracy of glaucoma.

Septiarini et al. [87], The objective of the suggested technique was to automatically segment the optic disc retinal fundus picture. I) creating an ROI image, ii) preprocessing, and iii) segmentation were the three primary procedures. The preprocessing process included normalization, scaling, and augmentation. To form the ground truth, the ophthalmologist’s labeled photos were transformed into binary images. The segmentation process required the ground truth to create the Unet[40] model’s structure.

A modified U-Net model for retinal vascular segmentation was proposed by K. Ren et al. (202)[77]. The data are initially preprocessed. Image enhancement is done via CLACHE, followed by the enhanced image slice. The process data is fed into the network using the U-Net architecture, and the BI-FPN fusion network is used to fuse the data. By combining the features at the top and bottom layers, the fused network increases the precision of vessel segmentation.

### 2.2. Statistical Features

One of the primary discriminatory measures of glaucoma infection is the cup-to-disc ratio, which is determined using segmented data. Furthermore, some statistical characteristics are computed, including kurtosis, rim entropy, and cup entropy. We now categorize three groups: i) Deep Learning Method; ii) Machine Learning; and iii) Graph-Based.

#### 2.2.1. Deep Learning Model

Bisento, Filho, Magalhaes (2020) [59] The suggested Generative Adversarial Network is linked to texture attribute-defined taxonomic diversity indices. Utilize GANs for OD segmentation to get better outcomes with fewer photos. When combined with taxonomic indexes, the GAN was made to retrieve textural characteristics from the divided OD area, which were essential for classifying glaucoma. To achieve 100% accuracy in ROC curve 1, they used a variety of classifiers, including Random Forest (RF), Sequential Minimal Optimization, and Multilayer Perceptron.

A comprehensive CAD system for automatically identifying and diagnosing glaucoma was proposed by Ferreira, Filho, Silva, et al. (2018) [47], who used deep learning, texture descriptors, and medical specialists’ knowledge as foundational data for tool construction. To train the CNN to segment the OD region, we use the ROI location that was supplied by an expert.CNN categorizes.

An evolving convolutional network (Ecnet), a non-handcrafted feature extraction technique, was proposed by Nayek, Das, Bhandary, and Acharya (2021) [70] for the automatic identification of glaucoma from fundus pictures. The extraction of discriminative features using multiple layers in this technique, including convolutional, compression, rectified linear unit (ReLU), and summation layers. Utilizing the Real Coded Genetic Algorithm (RCGA), the weights at various layers are optimized. Criteria that reduce intra-class variance and maximize inter-class distance are used to train the ECNet. We used different classifier techniques, and the SVM classifier technique gave the best results in diagnosing glaucoma. A dataset of 1426 fundus images—589 normal and 837 glaucoma—was used for the studies. The accuracy of the SVM model is 97.20 percent.

#### 2.2.2. Machine Learning

In comparison to traditional methods, some machine learning techniques have been researched and developed for automated glaucoma detection that can reliably identify glaucomatous damage on pathological tests and interpret retinal pictures quickly.

#### 2.2.3. Unsupervised Learning

Glaucoma detection may be possible with unsupervised learning, particularly when there is little labeled data or the objective is to find hidden patterns or groups in the data. Below, I’ll describe how unsupervised learning techniques can help with glaucoma identification.

Semmlow and Griffel (2021) [71], Local Binary Pattern (LBP), Gray Level Co-occurrence Matrix (GLCM), Gray Level Run Length Matrix (GLRM), Histogram Oriented Gradient (HOG), fuzzy-C means, and K-means are examples of statistical features that are used in unsupervised learning and also needed for the diagnosis of glaucoma.

Juneja, Minhas, et al. (2022) derived GLRM and GLCM characteristics from wavelet-filtered OCT images [71]. They employed correlation statistics, information gain, and gain ratio to identify discriminative features. Additionally, they used a 3D-CNN architecture for classification and feature extraction. Using SVM, K-NN, RF, and the probabilities from the 3D-CNN model, the final classification was combined using weighted decision fusion stratification and majority voting.

Karmawat, Gour, and Khanna (2019) [53] suggested a technique for creating a fundus image-based glaucoma diagnostic system. The optic cup is segmented using FCM clustering, and an elliptical curve fitting technique is utilized to provide smoothed borders. Glaucoma in fundus images from the Drishti GS1 collection is segmented, and characteristics based on global images are retrieved. To classify fundus images into normal and glaucoma classes using SVM and ensemble classifiers, the methods concentrate on optic cup segmentation utilizing fuzzy c means.

R. Shinde (2021) [72] suggested an offline computer-aided diagnostic (CAD) system that uses retinal fundus images to diagnose glaucoma. Deep learning, machine learning, and image processing techniques are used in this application. The brightest spot algorithm for Region of Interest (ROI) recognition and input picture validation is implemented using the Le-Net architecture. The U-Net architecture segments the optic disc and cup, while SVM, neural networks, and Adaboost classifiers are used for classification. LeNet model accuracy is 99%.

#### 2.2.4. Supervised Machine Learning

Parashar and Agarwal (2021) [73,74] suggested an SVM for glaucoma using the supervised algorithm of a machine-learning framework. A 2-dimensional variation mode decomposition method has been used in this investigation to obtain fundus images. Then, using the high-frequency modes, texture-based features such as the chip histogram and Zernike moment were calculated. The selected robust traits are subjected to the Student’s t-test methodology. To predict glaucoma, a multi-stage classifier (support vector machine) has been employed. A publicly accessible dataset is used to test the deployed technique’s efficacy. The accuracy of this model is 89.45%.

Raju, Shanmugam, et al. (2023) [86] Through temporal data construction using machine learning and logistic regression techniques, we created a predictive analytical framework for the early detection of glaucoma utilizing electronic health records (EHR) from more than 650 clinics and hospitals in the United States. The entire dataset was used to test four distinct machine-learning categorization techniques. Logistic regression models and five-fold cross-validation were used to calculate the accuracy, sensitivity, specificity, and F1 score. With an area under the receiver operating characteristics curve (AUC) score of 0.81 for glaucoma prediction one year before the onset of the disease, the XGBoost, multi-layer perceptron (MLP), and random forest (RF) performed similarly well to the logistic regression (LR) score of 0.73.

Tarcoveanu, Lean. Lisa et al. (2024) [94], Glaucoma is a complex, persistent, progressive, irreversible ocular neuropathy that mainly affects the elderly. In contrast to previous research in the literature that relies on picture collecting and processing, it is crucial to stress that the processed data are acquired from medical records. This method has the benefit of including many characteristics, which can show their potential influence, even though it is more difficult to manage. By employing the Neuro Solutions and PyTorch frameworks to build successive trials for almost ideal setups, artificial neural networks are utilized to examine the course of glaucoma. The PyTorch neural networks predicted the progression of glaucoma with over 98% accuracy, both in terms of statistical (visual field characteristics) and structural (retinal nerve fiber layer thickness) parameters.

#### 2.2.5. Graph-Based Approach

Tian, Zheng, Li, Du, and Xu (2020) [62],suggest an approach based on graph convolutional networks (GCNs) for optic disc and cup segmentation. The DRISTI-GS1 datasets were used in the recommended approach. The recommended approach consists of two primary components: The feature map of the input images is produced by the first component, a multi-scale convolutional neural network known as C-Net. The multi-scale feature is ignored by many conventional deep convolutional neural networks (DCNN), which restricts the ability to extract representative features. G-Net, a graph convolutional network, is the second component of the suggested approach. The multi-scale feature map from C-Net, concatenated with the original graph nodes and vertices, serves as the input for G-Net. We treat the segmentation task as a regression problem in our suggested segmentation method.

Jana, Thakur, Banerjee, and Mitra (2023)[89], provide a sophisticated graph-based approach that overcomes the shortcomings of traditional methods for automatically determining the precise location of the optic disc, By utilizing the K-dense sub-graph technique, our methodology provides a fresh viewpoint on optic disc localization. The damaged optic disc region and complex patterns in retinal pictures are identified. These models are 93% accurate.

### 2.3. Hybrid Feature

Combining structural and statistical information can greatly enhance the effectiveness of glaucoma diagnostic models. With statistical features that yield useful image-derived information and structural features that show clinical measures and anatomical changes in the retina, this method offers a thorough understanding of the situation.

Balasubramanian and Ananthamoorthy (2022) [31] suggested eliminating the fundus image’s characteristics by combining statistical and structural methods. A diagnostic procedure for glaucoma is based on the wrapper method that uses a kernel extreme learning machine (KELM) classifier and bio-inspired algorithms. Using a correlation-based feature selection (CFS) method, three feature subsets are produced from the pre-processed fundus pictures using bio-inspired algorithms. The salp swarm optimization-based KELM, which determines the ideal parameters of the KELM classifier network, is trained using the chosen features. The 7280-image public and private retinal fundus datasets to test the suggested methods. According to the experimental system, it may achieve a maximum overall accuracy of 99.61% with 100% specificity and 99.89% sensitivity.

Gung, Pen, Sung, Li, and Zhang (2022) [85] suggested a DSLN framework that performs better than the most advanced models and has strong generalization and robustness. It can identify glaucoma from multi-ethnic fundus pictures with accuracy and successfully overcome domain shift.

Thakur and Juneja (2020) [61], Thakur and Juneja present a novel collection of reduced hybrid features, derived from structural and neighboring aspects. This might be used as a second opinion by ophthalmologists. The cup disc ratio (CDR) and the disc damage likelihood scale (DDLS) are two of the structural features that were retrieved. Grey-level run length matrix (GLRM), grey-level co-occurrence matrix (GLCM), first-order statistics (FoS), higher-order spectra (HOS), higher-order cumulant (HOC), and wavelets are examples of non-structural characteristics. Using criteria like accuracy, specificity, precision, and sensitivity, the research concludes with a comparison of K closest neighbors (k-NN), neural networks (NN), random forests (RF), support vector machines (SVM), and naïve Bayes (NB).

The loss of picture spatial information when the convolutional character is input into fc layers is one of the main issues with CNN models for segmentation applications. FCN[18,23] is a network with 1x1 convolutions that carries out the functions of the fully connected layer without the need for any “Dense” layers. Variable inputs can be fed in when dense layers are absent. A few methods allow us to employ dense layers while preserving the dimensions of variable inputs. Convolutional and deconvolutional layers are used instead of the FC layers in the CNN architecture to maintain the original spatial resolution. FCN is frequently utilized in biological picture segmentation. Convolution and deconvolution are carried out end-to-end with multiple class classifications in FCN procedures.

A new collection of retinal fundus images known as Bangladesh Eye Hospital (BEH), which were classified as glaucomatous and healthy (nonglaucomatous) fundus images, was presented by M.T. Islam et al. [78]. The Bangladesh Eye Hospital in Dhaka (BEH) provided 634 color fundus photographs, of which 463 were normal/nonglaucomatous and 171 were glaucomatous. Two ophthalmologists thoroughly documented these images. From the clipped ocular cup and disc fundus images, we have employed four distinct deep learning algorithms—EfficientNet, MobileNet [34, 48], DenseNet [38], and GoogLeNet [12]—to detect glaucoma.

#### 2.3.1. Classification

These models perform well because they can employ extensive contextual data during training, which may reduce information loss and improve generalizability. According to their architectures, these models are divided into convolutional neural networks, autoencoder-based networks, attention-based networks, generative adversarial networks, geometric deep learning networks, and hybrid networks. This section examines research that focuses on these models for glaucoma diagnosis.

Deperlioglu et al. (2022) [82], Class Activation Mapping (CAM) was used to achieve XAI, enabling CNN to analyze images and provide interpretations based on heat maps. Twenty classification attempts were made using the Drishti-GS, ORIGA-Light, and HRF retinal image datasets to evaluate the hybrid solution’s performance. According to the performance evaluation, the ORIGA−Light dataset had the greatest mean values (accuracy: 93.5%, sensitivity/recall: 97.7%, specificity: 92.6%, precision: 93.8%, F1-Score: 95.7%, and AUC: 95.1%). Some physicians assessed the CAM-based XAI effect because the study’s XAI contribution included human analysis as well. The accuracy of the CAM-based XAI was 82.73%.

Yousefi, Pasquale, Boland, and Johnson (2022) [83], To find common patterns of VF loss, VFs were evaluated using an OHTS-certified VF reader and an unsupervised deep archetypal analysis program. Patterns of glaucoma damage that were discovered by machines were contrasted with those that were discovered by experts in the OHTS in 2003. Each eye’s longitudinal VFs were used to identify VF loss patterns that were highly correlated with the rapid advancement of glaucoma. Relationship between machine experts and the kinds of VF loss patterns linked to quick advancement. At conversion to glaucoma, the average VF mean deviation (MD) was -2.7 dB (SD = 2.4 dB), whereas the average MD of the eyes at the most recent appointment was -5.2 dB (SD = 5.5 dB). An MD rate of −1 dB/year or above was present in 50 out of 205 eyes.

Liao, Zou, Zhao, Chen, He, and Zhou (2019) [68], Three unique components are used in the proposed EAMNet to precisely identify local regions with specific traits and appearances to support the diagnosis of glaucoma. In particular, a well-designed CNN has been built to abstract hierarchical data for automated glaucoma diagnosis and semantic feature extraction. In addition to constructing an information passageway to close the gap between semantic and localization information at multiple scales and working with evidence activation, a novel technique called multi-layer average pooling (MLAP) is proposed to integrate features of various levels for an accurate glaucoma diagnosis. Both fully-supervised diagnosis and weakly-supervised evidence localization are produced by mapping this approach.

Hervella, Rouco, Novo, Ortega (2022) [84], A new multi-task method for segmenting the optic disc and cup and classifying glaucoma simultaneously. This method can enhance overall performance by using pixel-level and image-level labels during network training. Furthermore, pertinent biomarkers like the cup-to-disc ratio can be extracted thanks to the segmentation maps that are anticipated in conjunction with the diagnosis. The suggested methodology presents two technical innovations. First, a network architecture that increases the number of parameters shared by both jobs for simultaneous segmentation and classification. Secondly, by avoiding loss weighting hyperparameters, a multiadaptive optimization technique guarantees that both jobs contribute similarly to the parameter updates during training.

M.A. Majlan and his team [88] employed deep learning and computer vision techniques to diagnose glaucoma. We synthesize prior work in several architectural categories, including geometric deep learning, CNNs, attention networks, GANs, and autoencoders [21, 42]. Preprocessing includes grayscale transformation and adaptive histogram equalization (AHE) S. Das et al. [90]. An equalized picture is then subjected to the Frangi filter for vessel segmentation.

To predict the stage of glaucoma (mild, moderate, and severe) based on features often employed by ophthalmologists, J.N.K. et al. [91] suggest a version of the deep CNN model and validate it with those derived through visualization by CNN layers.

### 2.4. As per the Literature major Drawbacks are

- The way characteristics are used for classification varies depending on the study. Structural and statistical variables are used for classification in most of the research. The combined feature set has been utilized for classification in very few studies.
- In most methods, element choice has not been carried out using suitable ranking algorithms.
- Most researchers classify existing datasets, as real datasets used by some researchers.
- The technique of data augmentation.
- Multitask learning.
- Most researchers calculated the optic disc, optic cup, and CDR, but some researchers used retinal vessel segmentation.

### 2.5. The study aims

- Unbalanced Class.
- Model optimization for early illness detection
- Combining multimodal information.
- The classification of normal and abnormal retinal pictures, a reduced significant unique feature set comprising structural and non-structural features, has been presented.

### 2.6. Novelties and Contributions

Our suggested model substantially contributes to the area and adds several new features. Below, we describe the novelties and contributions of the suggested Miniunet.

Novelties:

- Using the DRIVE, STARE, and Dristi-GS1 data sets, we must present a novel MiniUNet model to minimize the layer while sacrificing the greatest accuracy.
- Using this model, We segmented the optic disc and retina vasculature from the fundus image.

Contribution:

This model included AttnGatingBlock, UnetGatingSignal, and UnetConv2Dpro, which are used in the deep learning model and glaucoma detection.

## 3. PROPOSED METHOD

Image segmentation in Deep Learning used U-Net[16], Dense Convolution[38], BConnvLSTM[42], and MobileNet[34], We proposed MiniUnet shown in the figure.

### 3.1. ENCODING PATH

There are four stages in the Mini_UNet’s encoding path. The 2D convolution 3x3 filter is followed by a Rectified Linear Unit (ReLU) and a 2x2 Maxpool function [6]. With each step, there are fewer feature maps. Layer by layer, the encoding path expands the demonstration’s dimension while progressively eliminating picture demonstration. The image demonstration’s encoding path, high dimension, and raised semantic information make up the final layer. The main U-Net architecture had more convolution layers and a last-step encoding path. The main U-Net network might be trained on superfluous features in the consecutive convolution. Densely connected [38] convolution layer moderate U-Net Problem. This technique helps enhance the performance, reprocessing the feature map throughout the entire network. Feature maps are trained on the previous convolution layer, the existing layer is merged, and then the next convolution layer is forwarded. The regular convolution layer has many problems in resolving the dense convolution layer. It helps dense networks remove redundant features and then learns a new set of features. This technique improves the performance of the entire network. Densely connected networks avoid the danger of gradient decline or explosion. The backward path sends the gradient faster to its specific location in this network. The last phase in the N block’s [51] convolutional layer sequence is encoding, as can be seen in the image.

### 3.2. DECODING PATH

The feature mappings from the narrowing path are copied and cropped to the decoding path in the original U-Net. The output of the upsampling function is merged with these feature maps. The mini-Unet uses two kinds of feature maps.

### 3.3. BATCH NORMALIZATION

Up-sampling function denoted 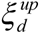 passed in Batch Normalization function and produced 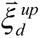 . Since every step has been adjusted to fit the new distribution, the training process is incredibly slow. It is used to make a neural network more powerful. Batch Normalization[17] is a technique that artificially speeds up a neural network’s training process.

### 3.4. TRAINING PROCESS

The cross-entropy loss function is combined with the final feature map that was determined using a pixel-wise soft-max[16] for energy. The training data set precomputes the weight map for every ground truth to reimburse various pixel frequencies. This network starts between two distinct cells and learns the bounds of minor partitions.

The morphological procedures utilized to compute the weight map equation and divide the border area, The equation is

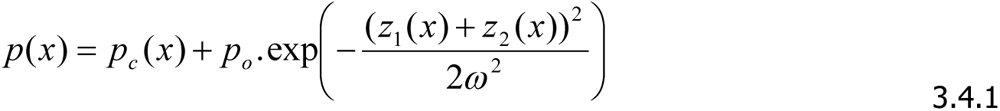

Where 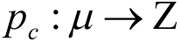 denotes weight class occurrence and 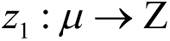 denotes distance the resident of the adjacent cell and 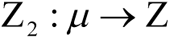 denotes distance resident of the second adjacent cell.

Despite its high activation, this network’s usage of many convolution layers and a variety of routes yields decent results. Our approach is based on the ReLU activation model and the Gaussian distribution. Each 64-component feature vector and 1x1 convolution layer be used.

### 3.5. DATABASE DESCRIPTION

Every experiment uses the Digital Retinal Images for Vessel Extraction (DRIVE) database for training and evaluation. 40 RGB fundus photos, each measuring 565 × 584 pixels, make up DRIVE. Every image comes with Field of View (FOV) masks and manual labels. The database is split equally between a testing set and a training set. For validation, a subset of four photos is chosen at random from the training set. A preprocessing pipeline is used to prepare the raw images, extract the green channels, balance the inhomogeneous lighting with CLAHE, and standardize the pixel intensities within the FOV masks to (-1, 1). To eliminate possible border effects and guarantee meaningful comparison, the edges of every FOV mask are degraded by four pixels. Furthermore, to highlight thin vessels, multiplicative pixel-wise weight maps w are created from the manual labels using the formula: 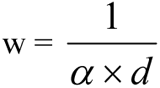, where d is the vessel diameter in the provided manual label and α is manually adjusted to 0.18.

### 3.6. DATA EXPANSION

A segmentation network with a very thin gloss image seems to be trained using the input concept of very random flexible deformation in the training data. Using random displacement vectors on standard 3x3 grids, we produce gentle deformation. Per-pixel displacements were calculated using the bi-cubic interpolation method. Further inferred data expansion is carried out by drop-out[9] layers at the end of the narrowing path.

## 4. ARCHITECTURE

**FIGURE 3.1.**
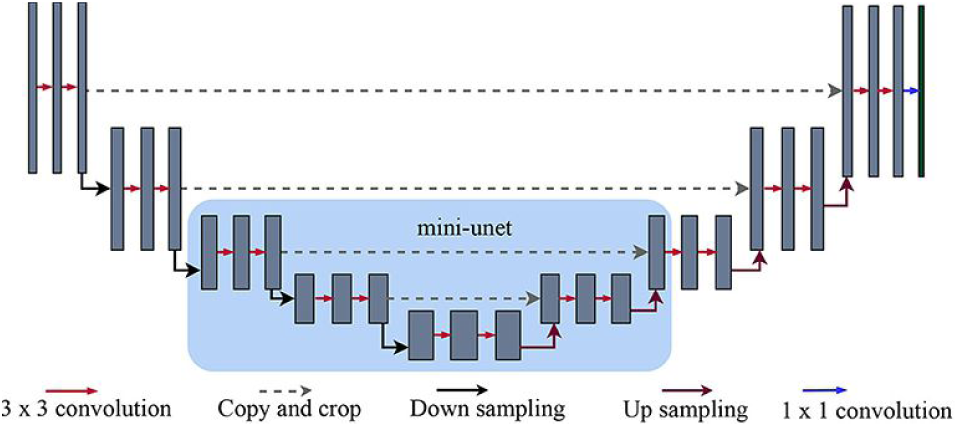
Mini-Unet architecture and densely connected convolution

Mini-Unet is an architecture for semantic segmentation. The encoding path and the decoding path are its two components. Level minimization is this architecture’s main objective. Figure 3.1 illustrates where to minimize the level, This design changed Version U_Net. The encoding path adheres to a convolutional network’s standard topology. ReLU and a 2x2 max pooling operation with stride 2 are used for the down-sampling after two 3x3 unpadded convolutions have been applied repeatedly. As more and more picture demonstrations are eliminated, the encoding approach progressively increases the demonstration’s dimension. Both the cropped feature map from the encoding path and the decoding path feature map are followed by 3x3 convolutions and ReLU, respectively, and 2x2 up-convolutions. Each of the 64 components in the final layer uses 1x1 convolution. This design finds the optic disc and removes the retinal vessel.

**FIGURE 3.2.**
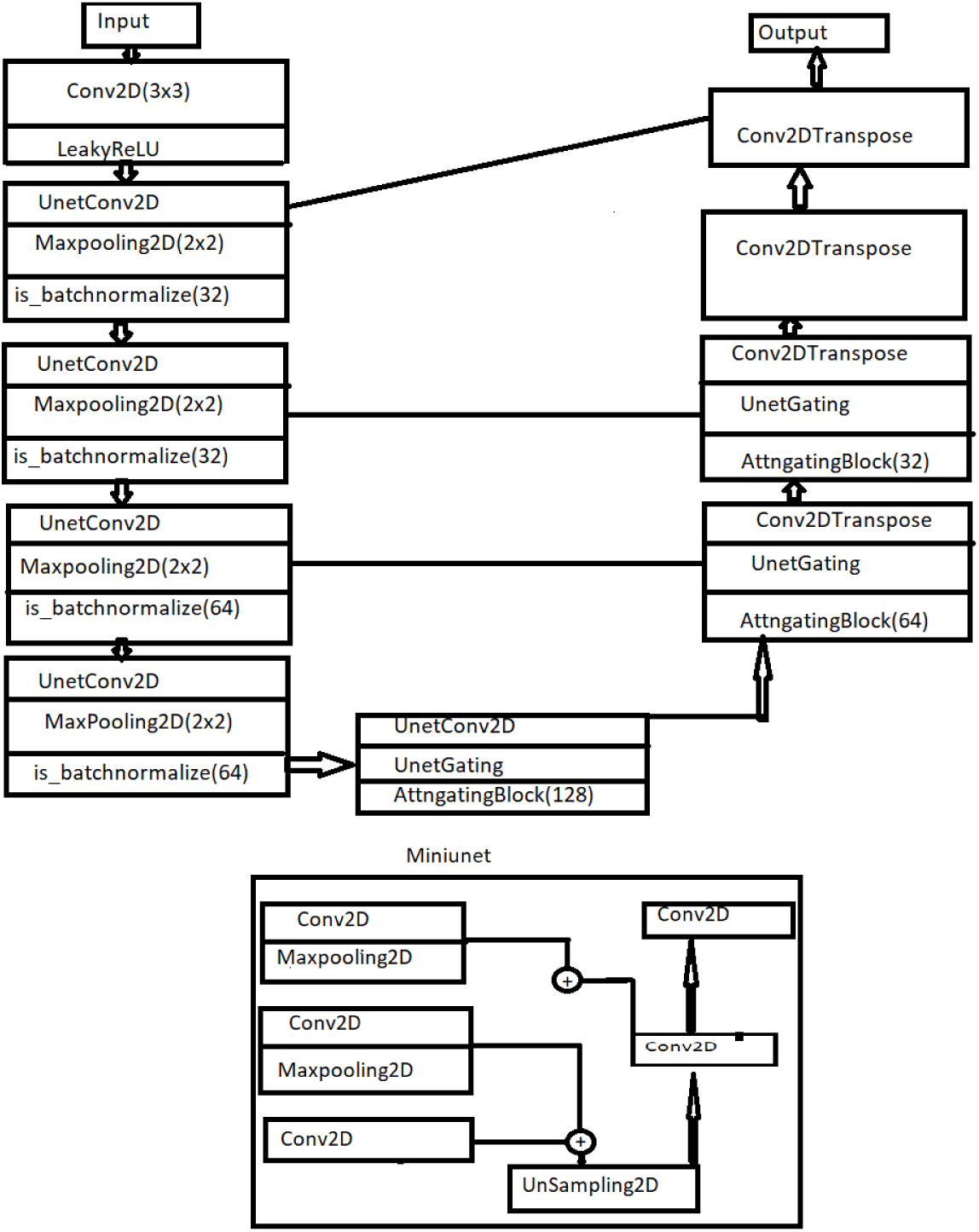
Graphically Demonstrate of the proposed Mini-Unet Model.

## 5. Proposed Miniunet Algorithm

1. Data Preparation: Data augmentation and cropping of fundus images. The Mini-Unet model was proposed to determine the location of the optic disc and segment the vessel. We used both manual and masked photos as input images in our investigation. The input image is 565x584x3, with channel 3 and dimensions of 565 by 584. To eliminate the impact of illumination variance throughout the image, each cropped fundus image is normalized with a resolution of 512 by 512, and the mean value over all the pixels is subtracted from each pixel to prevent the data from being overfitting. Also, we used a threshold value; the threshold value ranges between 0 and 1.
2. Training algorithm: our fundus picture representation’s thorough training and testing process. There are four components to our algorithm: Data preparation, initialization, and training in representation are the first three steps. Three highly connected layers make up MINI_UNET, our algorithm, and two of those layers are concatenated. The 2D convolutional layer, the 3 by 3 kernel size, and the Relu activation function are the first three layers. Concatenate the final result with the first convolutional layer after concatenating the final two layers. We employed the sigmoid activation function in the final concatenation step. To optimize weight, we trained our network using SGD and backpropagation approaches. Our method requires relatively less time than other existing techniques since it requires fewer layers, which means more time. After training, the pass can be forwarded to receive the representation results. Our binary descriptor has a bit length of 1024. To separate the optic disc and retinal vasculature, we employed a threshold of .5. There were a lot of tiny pixels, so take one out.

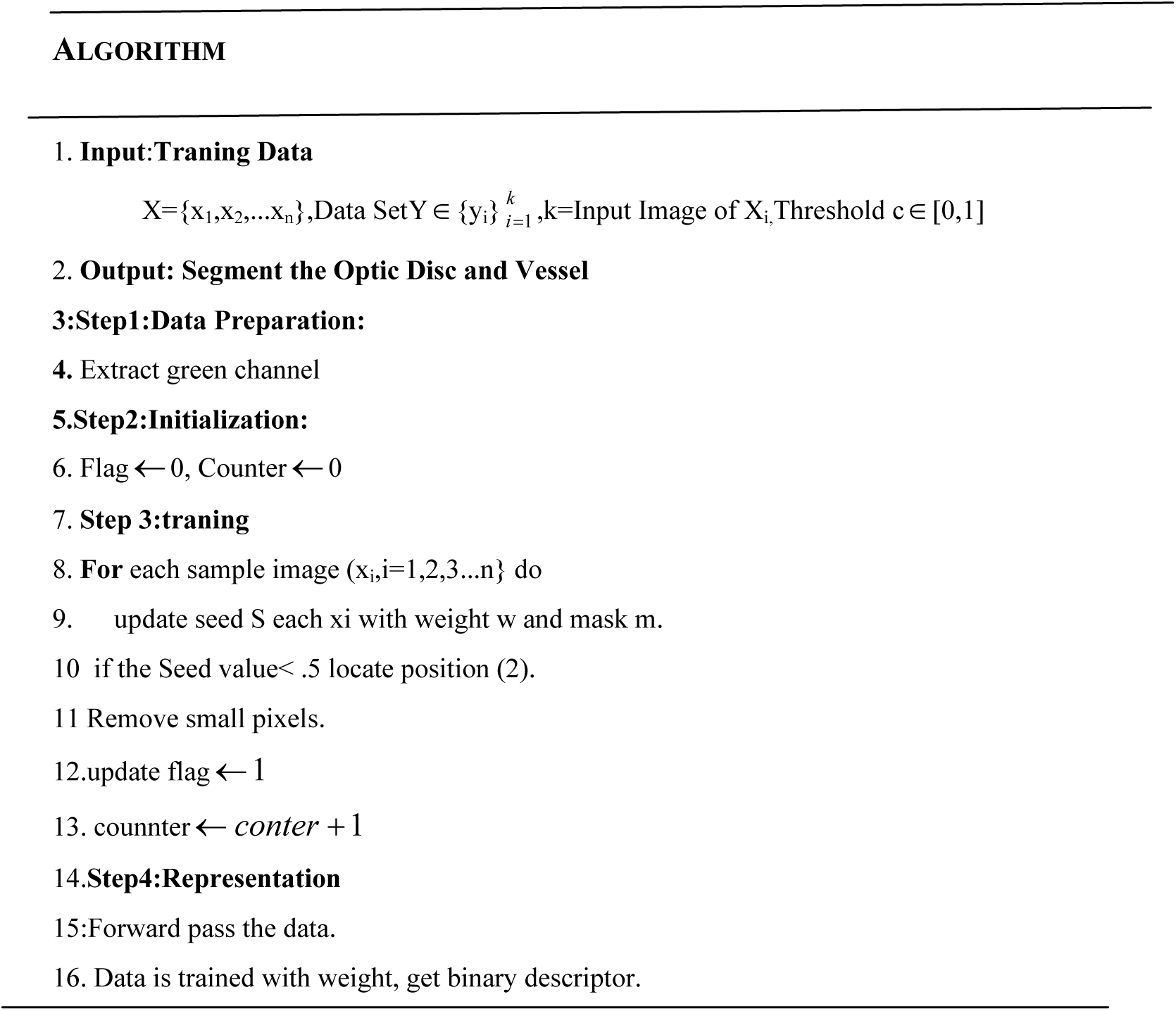

## 6. EXPERIMENT RESULT

**FIGURE 6.1.**
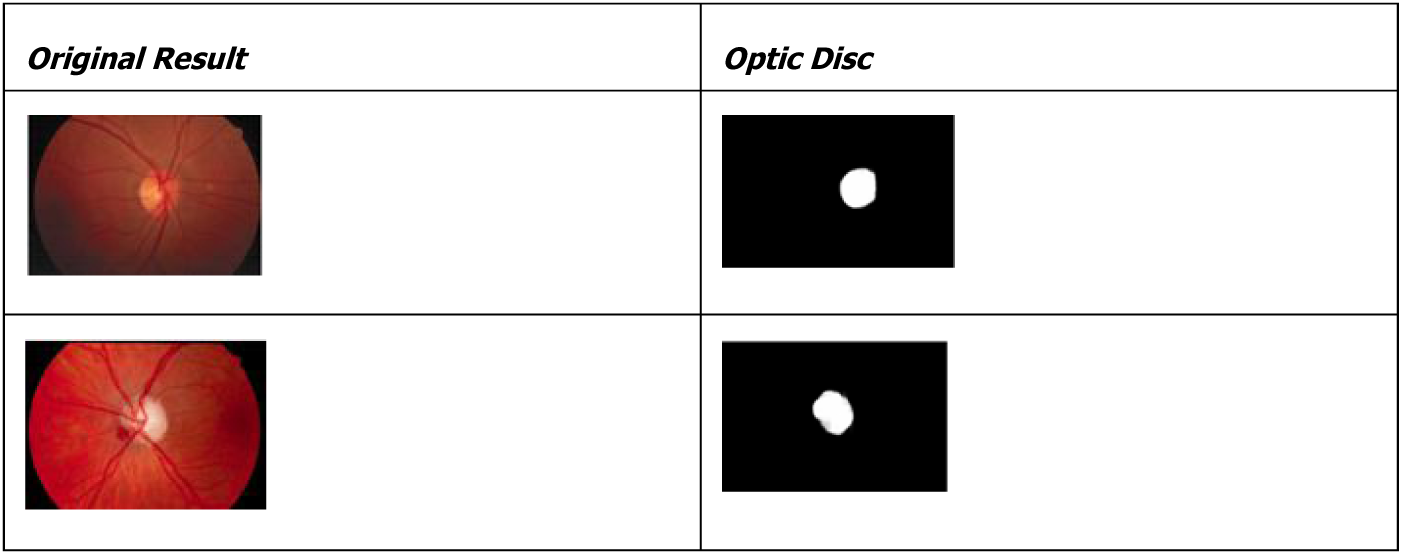
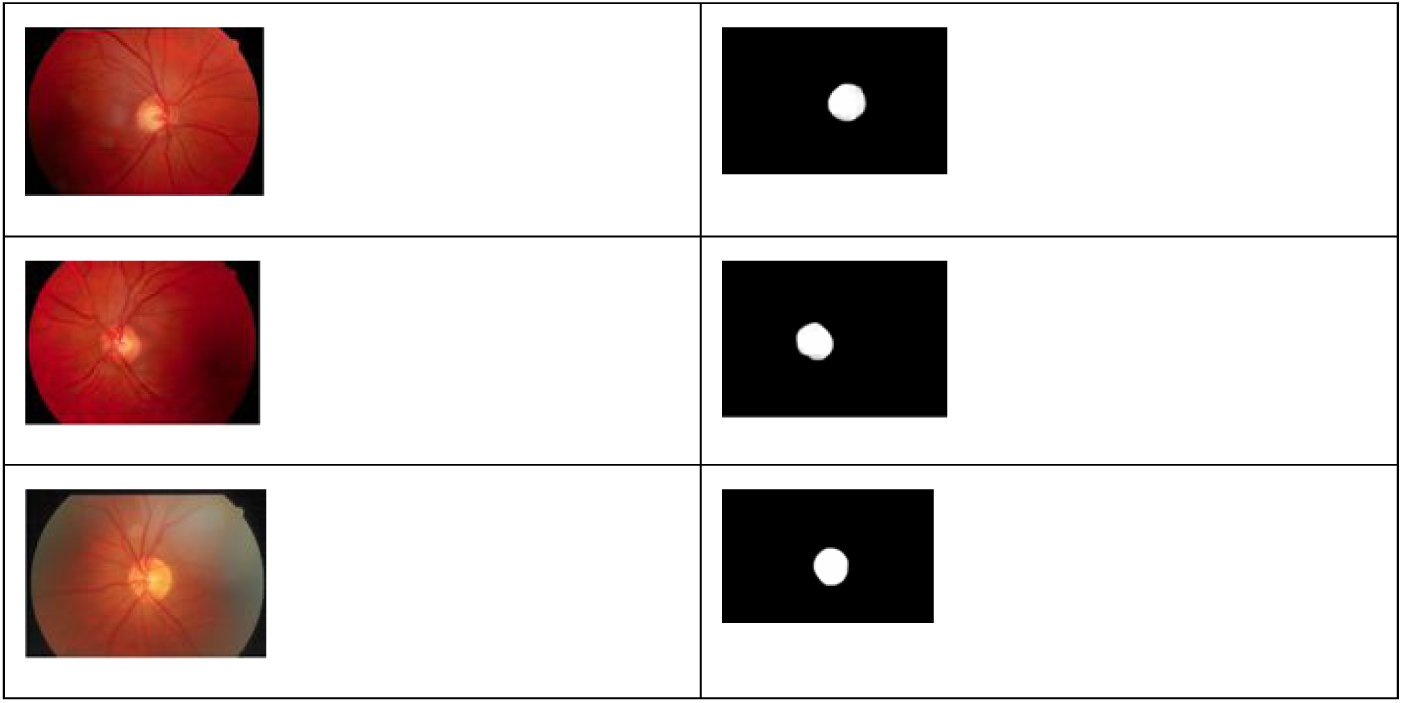
Optic Disc Segmentation Using Proposed Mini-Unet Model

**FIGURE 6.2.**
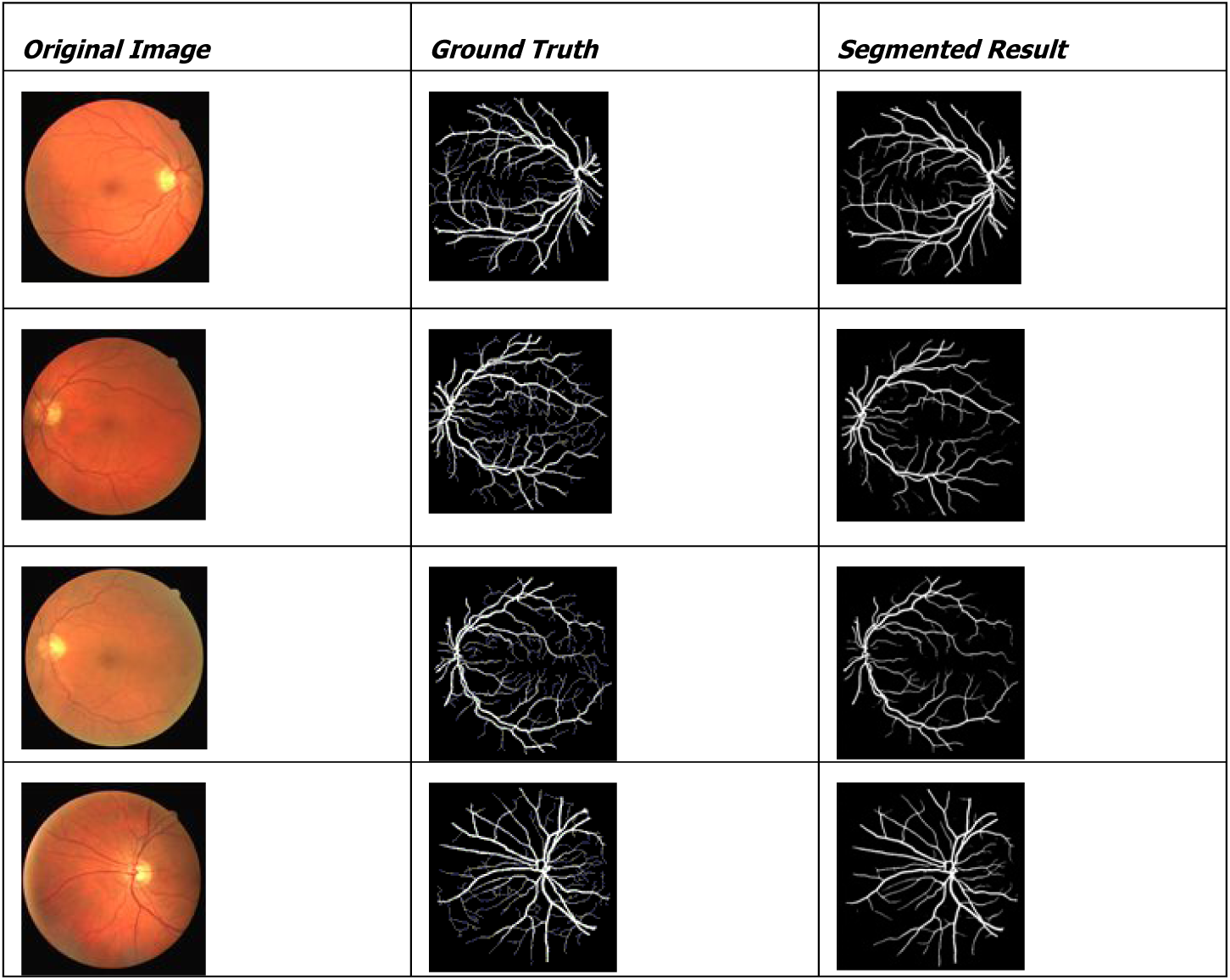
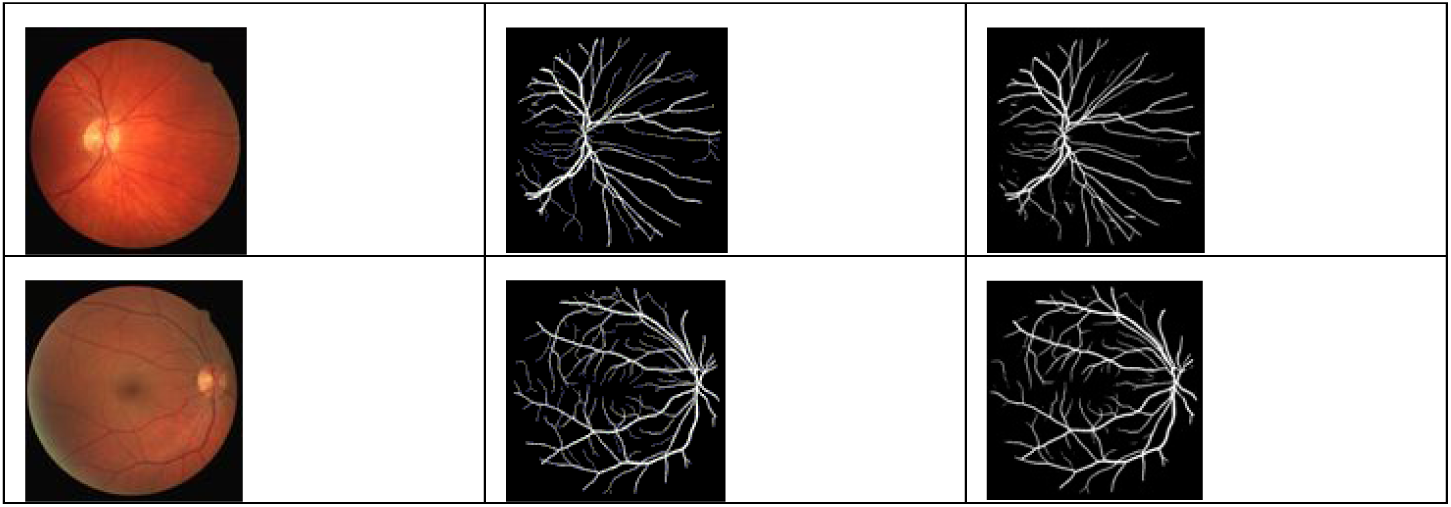
Retina Vessel Segmentation Using Proposed Mini-Unet Model

We test mini_unet on public standard data sets segmented by DRIVE, STARE, and CHASEDB. Every data set is used for optic disc localization and retinal vascular segmentation. TensorFlow and Keras are implemented in the mini-UNet backdrop. For the experimental comparison, we compute performance metrics such as precision, F1-score, Jaccard similarity score, accuracy (AC), sensitivity (SE), specificity (SP), area under the ROC curve, and area under the precision-recall curve. We use 20 sample photos for training and another 20 sample images for testing out of the 40 color retinal images in the Drive dataset. The photos have an input size of 565 x 584. Some samples are not enough for deep neural network training. Initially, patches are randomly divided into input images. Twenty training images consist of approximately 182,000 patches, of which 160,000 are used for training and the remaining 22,000 for validation. At first, our approach used batch sizes of 16 and epochs of 30. The two primary components of the loss function in this study are weighted loss and L2 norm with regularization. An SGD optimizer with a decaying learning rate set to 5x10^-5^ is used to minimize the objective function. Based on the validation loss, early stopping is used. 50,168 168x168 patch-sized photos are included in each batch, and data augmentation methods like rotation, shearing, additive Gaussian noise, and intensity shifting are used. Figure 6.1 displays the original color image in the first column; the second column shows the optic disc and FIGURE 6.2 Finds the retinal vessel segmentation; the first column is the original image, the second column is the ground truth image, and the third column is the segmented image. The mini-UNet model architecture was employed in two segmentations. Retinal vascular segmentation using the Maxflow [56]-based technique, although the results are not particularly noticeable. The various models and the quantitative outcomes of the suggested model are listed in Table 1.

**TABLE 1.**
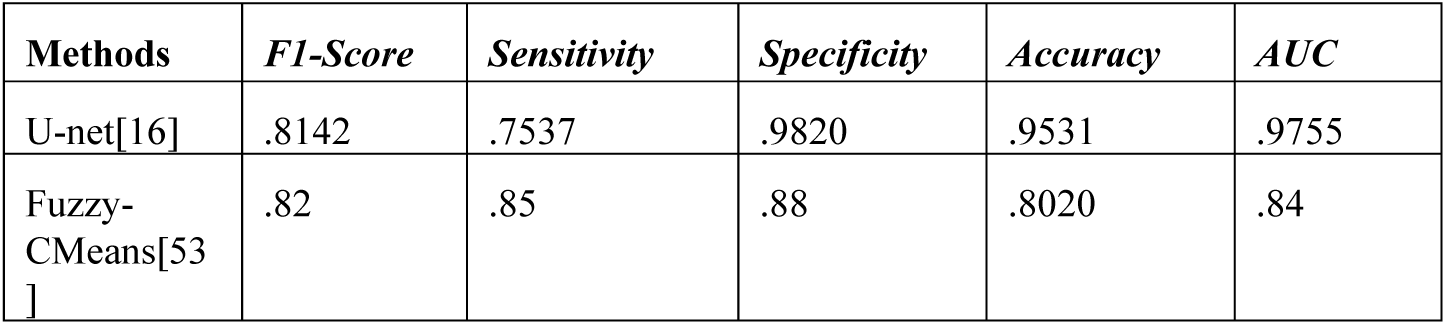
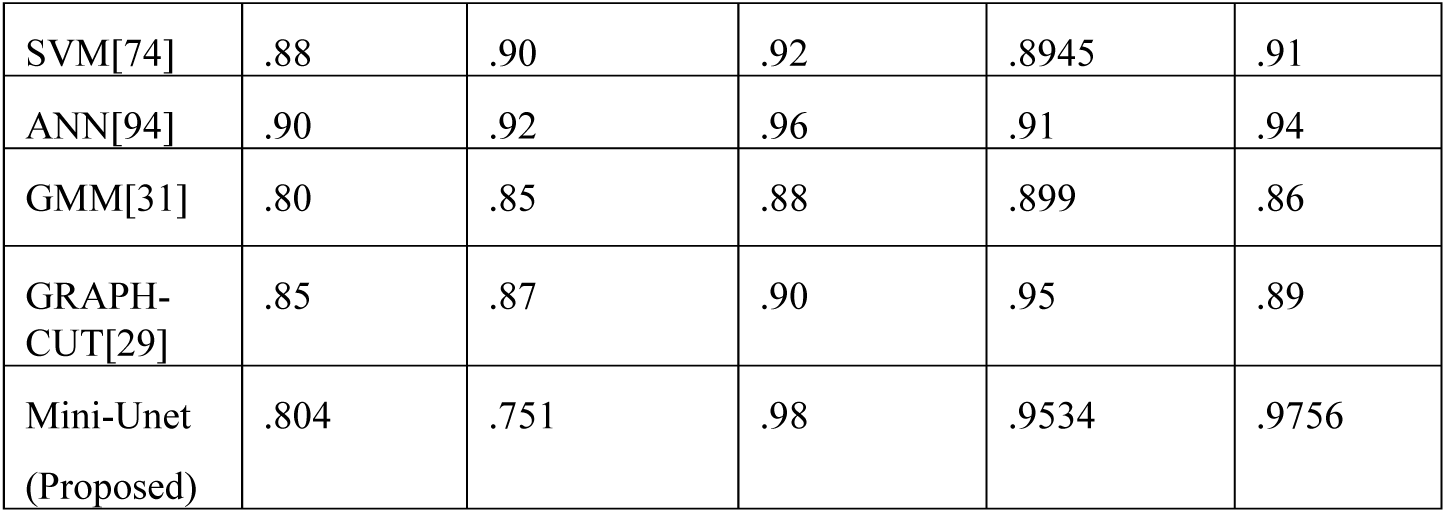
Performance Comparison Mini-Unet(Proposed) and Existing Model.

**FIGURE 6.3.**
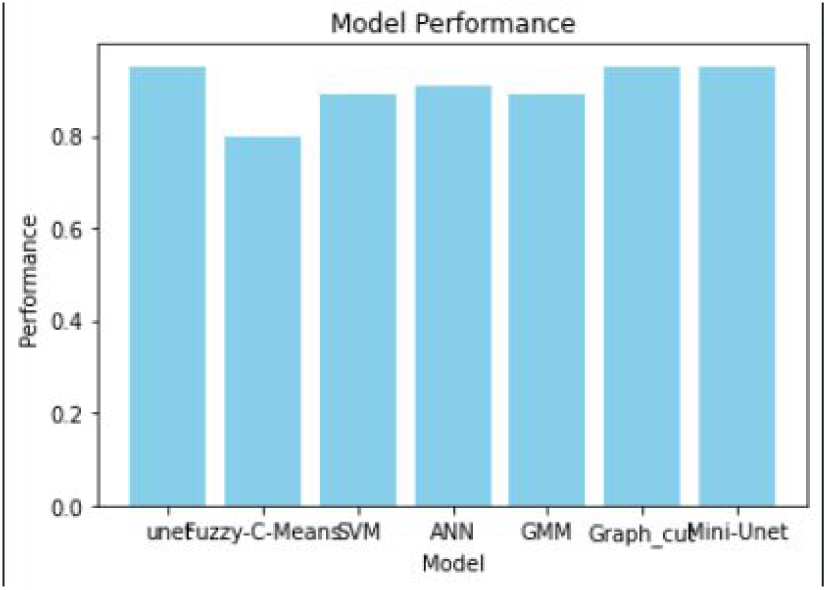
Graphically represent Different model with Accuracy

The Receiver Operating Characteristic(ROC) curves in Figures 6.4 and 6.5 show the proposed model’s overall performance. The ROC curve, which plots the true positive rate against the false positive rate, represents the true positive rate (TPR) and false positive rate (FPR).

**FIGURE 6.4.**
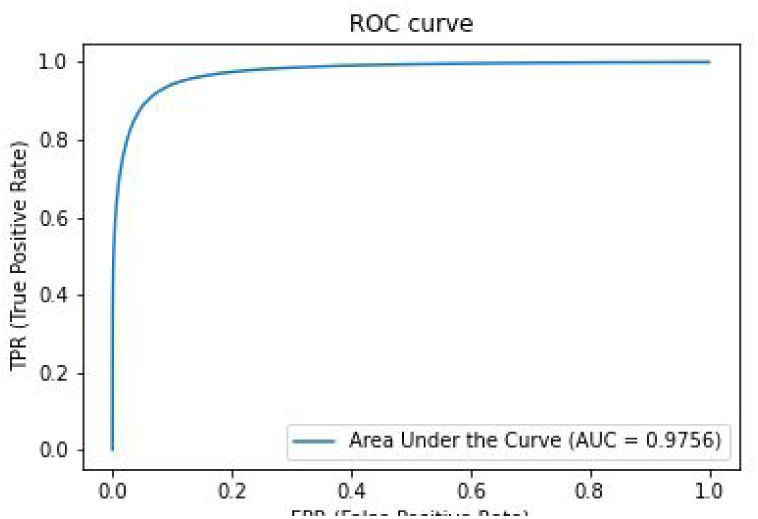
Graph of FPR VS TPR ROC curve Proposed Mini-Unet Model

**FIGURE 6.5.**
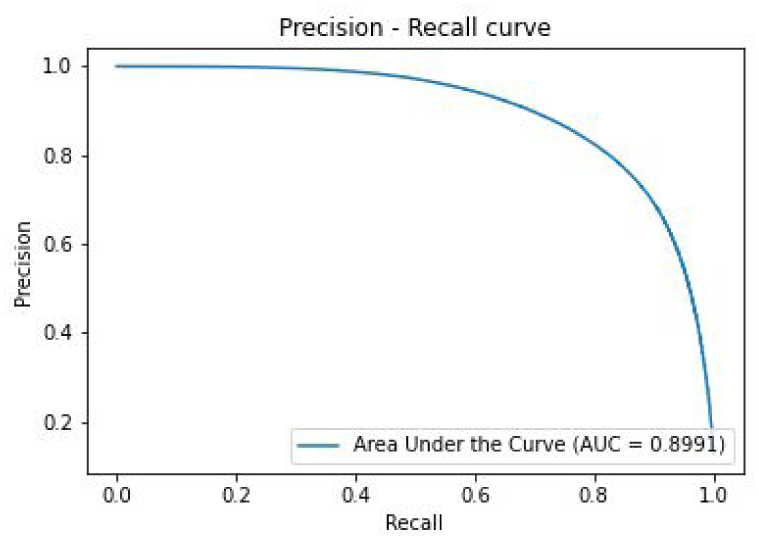
Graph of Recall vs Precision Precision call Proposed Mini-Unet

TABLE 2. Compares Structural Feature with feature, classification, Data Type, Accuracy given below:

**TABLE 2:**
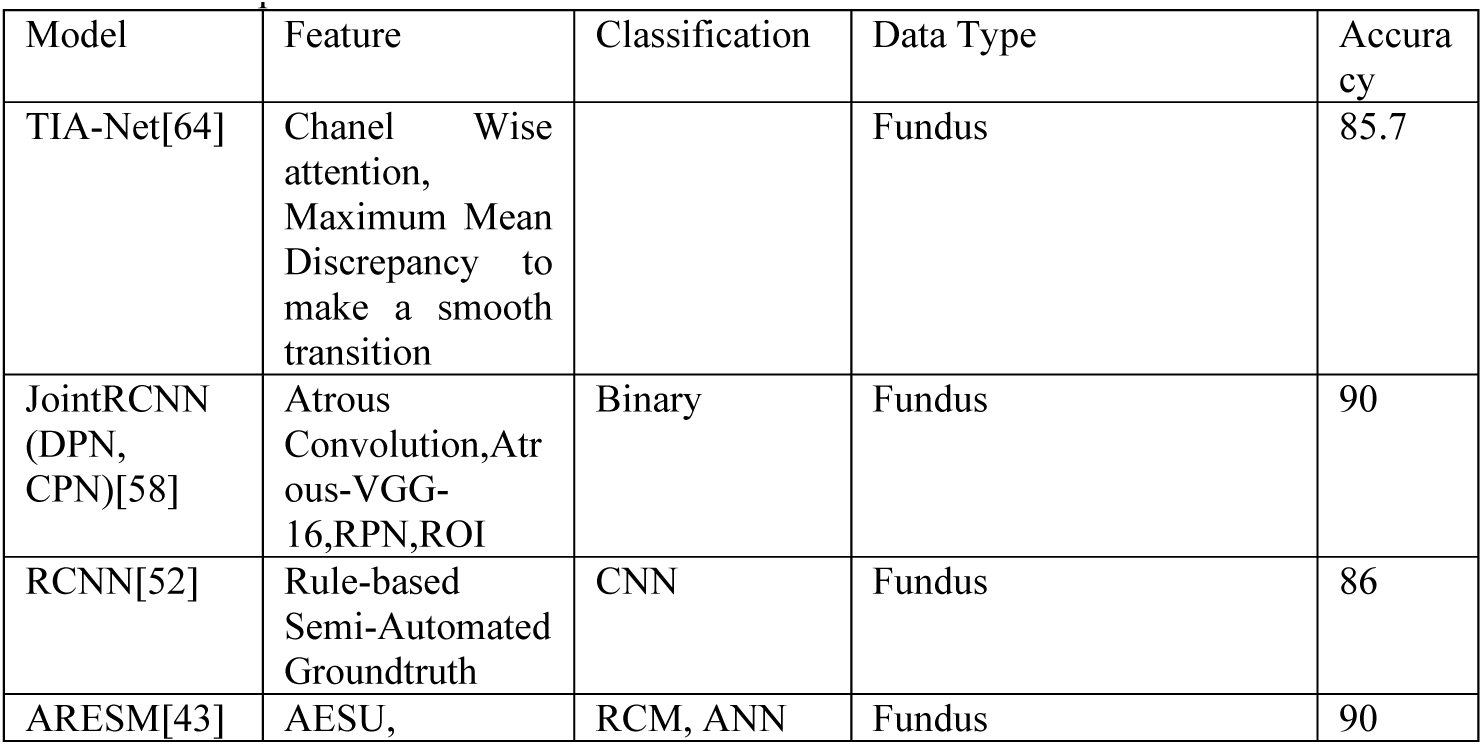
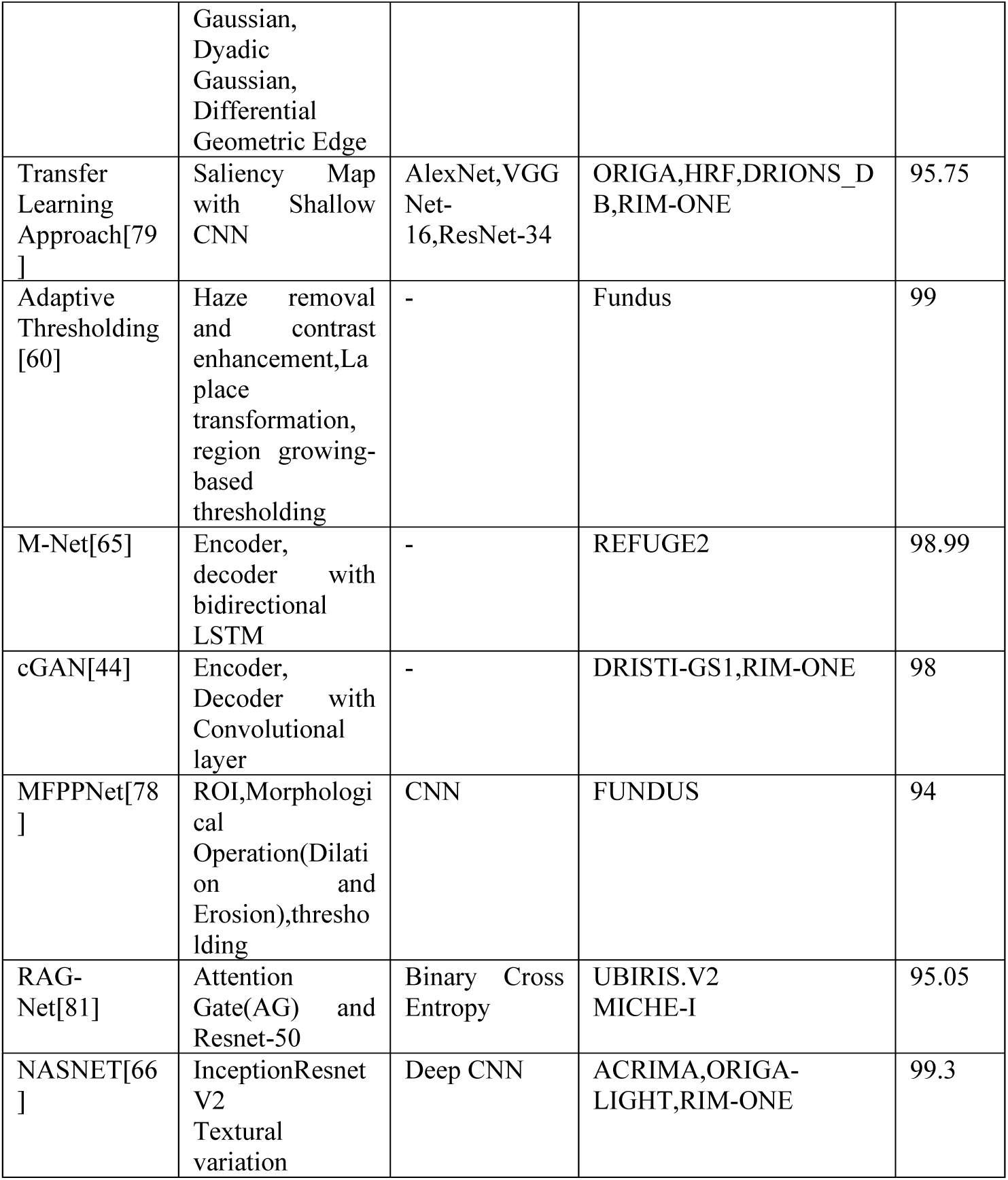
Comparison Structural Feature Extraction Different Model.

TABLE 3. Compares Statistical Features in different models with feature, classification, Data Type, Accuracy given below:

**Table 3:**
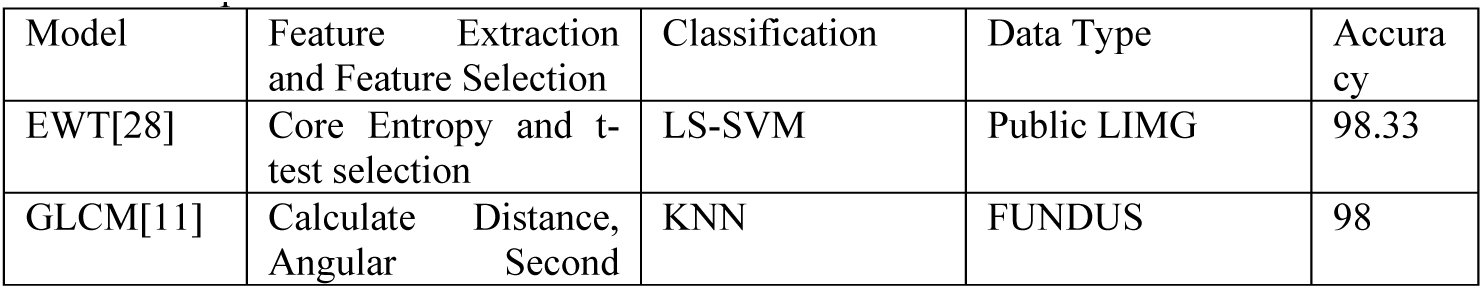
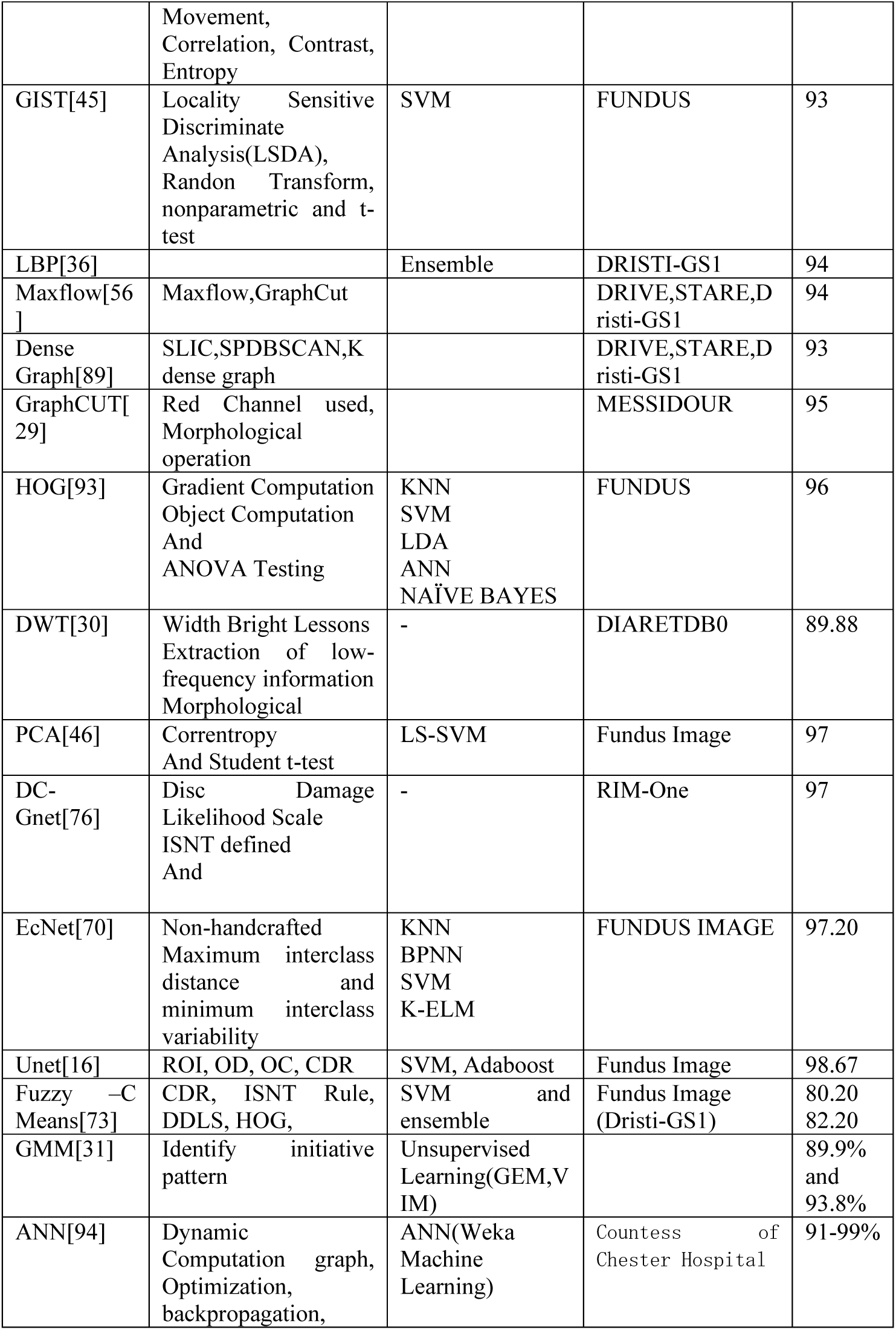
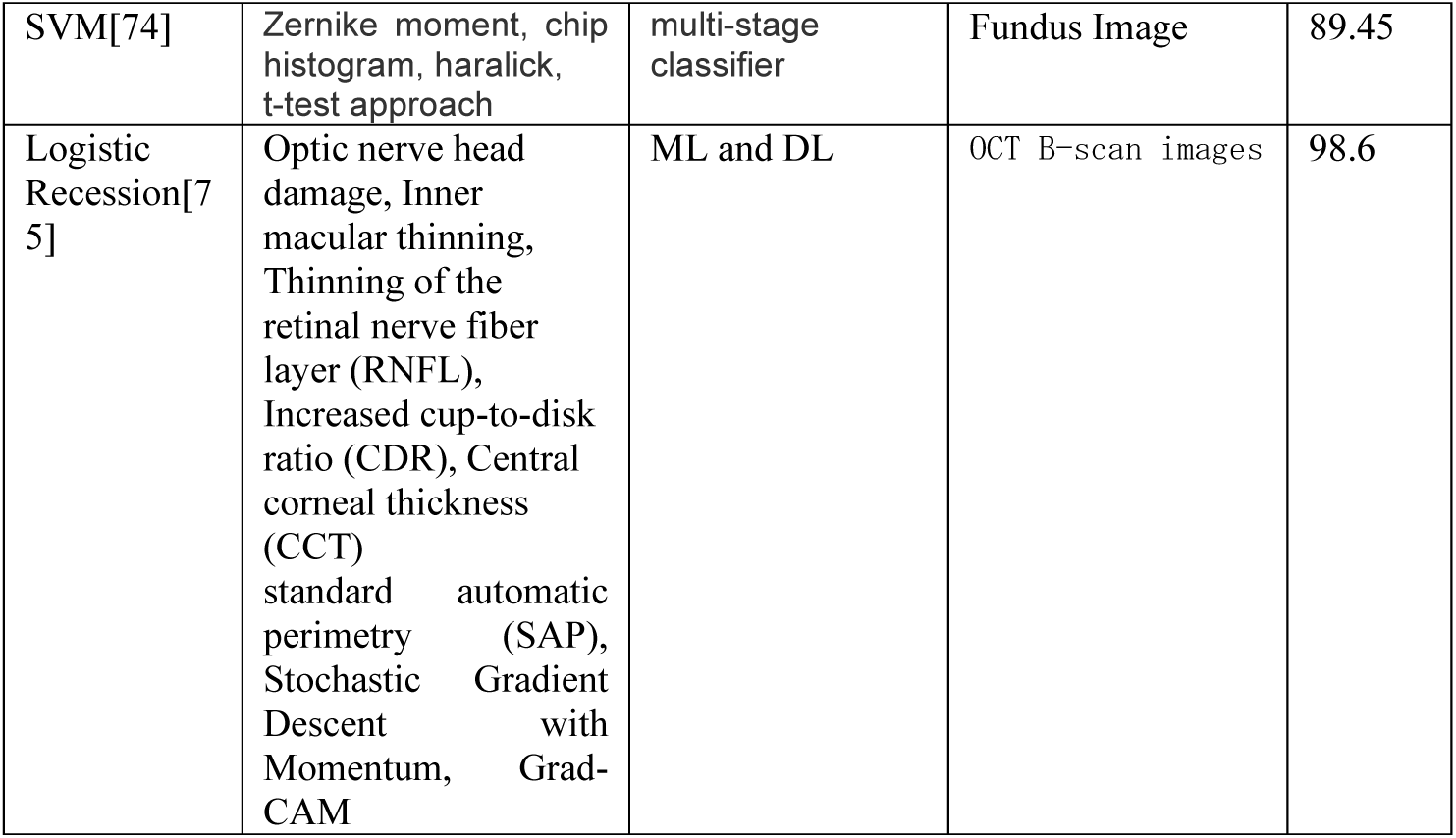
Comparison Statical Feature Extraction Different Model.

TABLE 4. Compares Hybrid Feature with a different models with the feature, classification, Data Type, Accuracy given below:

**Table 4:** Comparison Hybrid feature extraction different model.

| Model | Feature Extraction and Feature Selection | Classification | Data Type | Accuracy |
| --- | --- | --- | --- | --- |
| Fuzzy-C Means[53] | CDR, ISNT Rule, DDLS, HOG, | SVM and ensemble | Fundus Image (Dristi-GS1) | 80.20<br>82.20 |
| Wrapper[61] | Disc damage likelihood scale, Cup to disc ratio, GLRM, GLCM, FOS, HOS, | SVM, PCA | Fundus Image |  |
| MobilenetV2[95] | Enhance Segmentation Accuracy, Fine Tuned, GAN | MobilenetV2 | RIM-ONE(Fundus Image) | 98.9 |

## 7. CONCLUSION AND FUTURE WORK

We have developed a hybrid method for detecting optic disc and retina vessel segmentation in images. This method minimizes the layer structure U-Net model. We compared to existing method and our proposed method. Our proposed method has better results than other existing techniques and more accurate results. Looking ahead, Our study is being conducted in two primary directions. We are eager to investigate how CNNs[80] could enhance our detection technique, and we first intend to test our approach for noisy photos. This is important for real-world applications since it will help us understand how well our strategy performs in less-than-ideal settings. Our overall objective is to use the hybrid model to identify thin vessels, locate optic discs and cups accurately, and solve issues caused by noise in images.

## Data Availability

existing data

https://paperswithcode.com/dataset/drive

## ACKNOWLEDGMENT

I am grateful for my guide’s assistance and my efforts.

## Biographies

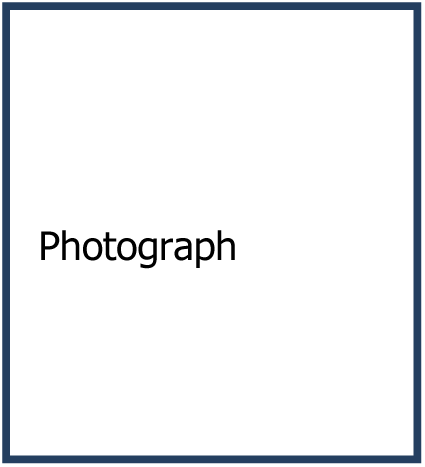

Author Name

